# Association of gamma-glutamyl transferase variability with risk of osteoporotic fractures: A nationwide cohort study

**DOI:** 10.1101/2022.10.28.22281658

**Authors:** Dongyeop Kim, Jee Hyun Kim, Heajung Lee, Iksun Hong, Yoonkyung Chang, Tae-Jin Song

## Abstract

**Objectives:** Gamma-glutamyl transferase (GGT) is related to inflammation, osteoporosis, and vascular diseases. Recently, changes in metabolic parameters have been proposed as disease biomarkers. We aimed to assess longitudinally the association of GGT variability with osteoporotic fractures.

**Methods:** From the National Health Insurance Service-Health Screening Cohort database, participants who underwent three or more health examinations between 2003 and 2008 were included (n=1,072,432). Variability indexes were as follows: (1) coefficient of variation (CV), (2) standard deviation (SD), and (3) variability independent of the mean (VIM). The primary outcome was occurrence of osteoporotic fracture, defined as identification of one of the following international classification of diseases-10 codes: vertebral fracture (S22.0, S22.1, S32.0, S32.7, T08, M48.4, M48.5, M49.5), hip fracture (S72.0, S72.1), distal radius fracture (S52.5, S52.6), or humerus fracture (S42.2, S42.3).

**Results:** During a median of 12.3 years (interquartile range 12.1 – 12.6), osteoporotic fractures occurred in 49,677 (4.6%) participants. In multivariable analysis, GGT variability based on CV positively correlated with the occurrence of osteoporotic fracture (adjusted hazard ratio [HR] of the highest quartile compared with the lowest quartile 1.15, 95% confidence interval [CI] 1.12-1.18, *P* < 0.001). These results were consistent even when GGT variability was defined by SD (adjusted HR 1.22, 95% CI 1.19-1.25, *P* < 0.001) and VIM (adjusted HR 1.12, 95% CI 1.09-1.15, *P* < 0.001).

**Conclusions:** Increased GGT variability is associated with an increased risk of osteoporotic fractures in the Korean population. Maintaining constant and stable GGT level may help reduce the risk of osteoporotic fractures.

## Introduction

Gamma-glutamyl transferase (GGT) is one of the representative biomarkers of liver disease. Recently, it has been reported that GGT level is also associated with other diseases including cardiovascular disease and osteoporosis and decreased bone mineral density (BMD) and with mortality as well.(1–3) To confirm the association between a specific biomarker and disease risk, it may be more reliable to determine whether variability in multiple measurements is more highly associated with risk than is change in only one measurement. In this context, GGT variability is closely related to risk of myocardial infarction, stroke, and heart failure.(4,5)

Osteoporotic fractures are a common but serious health problem in humans. In particular, the incidence of osteoporotic fractures increases with age and explosively increased with the transition to an aging society worldwide.(6) Osteoporotic fractures can cause disability due to the disease itself, and the accompanying economic burden is a significant social problem.(7) Among the various sites of osteoporotic fractures, hip and spine fractures are among the most severe and can cause mortality.(8) Risk factors of osteoporotic fracture include low body weight, premature menopause, smoking, alcohol, physical inactivity, steroid use, and vitamin D insufficiency. In this setting, information on correctable or preventable risk factors for osteoporotic fractures is needed.(9)

In previous studies, increased blood GGT level has been associated with osteoporosis and decreased BMD.(3,10) These results support the possibility that GGT is involved in bone metabolism and may be associated with osteoporosis and related fractures. Moreover, variability in metabolic parameters, including GGT, is closely related to inflammation-associated metabolism and dysregulation of homeostasis, which may be associated with cardiovascular diseases and osteoporosis.(5,11) However, the association between long-term variation in GGT and osteoporotic fractures remains to be elucidated. Our hypothesis is that increased GGT variability is associated with increased risk of osteoporotic fractures, and we used a nationwide cohort database with a longitudinal setting to investigate the association between GGT variability and osteoporotic fractures.

## Methods

### Data source

The National Health Insurance Service (NHIS) is a single insurer managed by the Korean government, covering nearly 97% of the population, with the remaining 3% covered by the Medical Aid program.(12) The NHIS subscribers are encouraged to undergo a standardized health checkup every year, from which a national health screening database has been established (NHIS-HEALS). The NIHS-HEALS cohort consists of a stratified random sample representing approximately 1,236,000 people between the ages of 40-79 years who underwent health screenings, representing 25% of the total population (dataset number: NHIS-2021-01-715).(13) This database contains information on the individual demographics, socioeconomic status, medical examination information including laboratory test results, and claims information including diagnosis, prescription, and treatment modalities. Demographic data include basic information such as height and weight and surveys about lifestyle, including smoking and alcohol history. The Institutional Review Board of Ewha Womans University College of Medicine (SEUMC 2022–07–053) approved this study with a waiver of informed consent because the study used deidentified retrospective cohort data.

### Study population

Participants from the NHIS-HEALS database who underwent health checkups between 2003 and 2008 were included (n=1,236,589). Those with missing data on at least one variable of interest (n=91,251) were excluded. Moreover, participants with a previous history of osteoporotic fracture from January 2002 to the date of checkup (n=18,823) and participants with fewer than three repeated GGT measurements (n=54,083) were also excluded. Finally, the data from 1,072,432 people were included and analyzed in this study (Fig 1).

**Fig 1.**
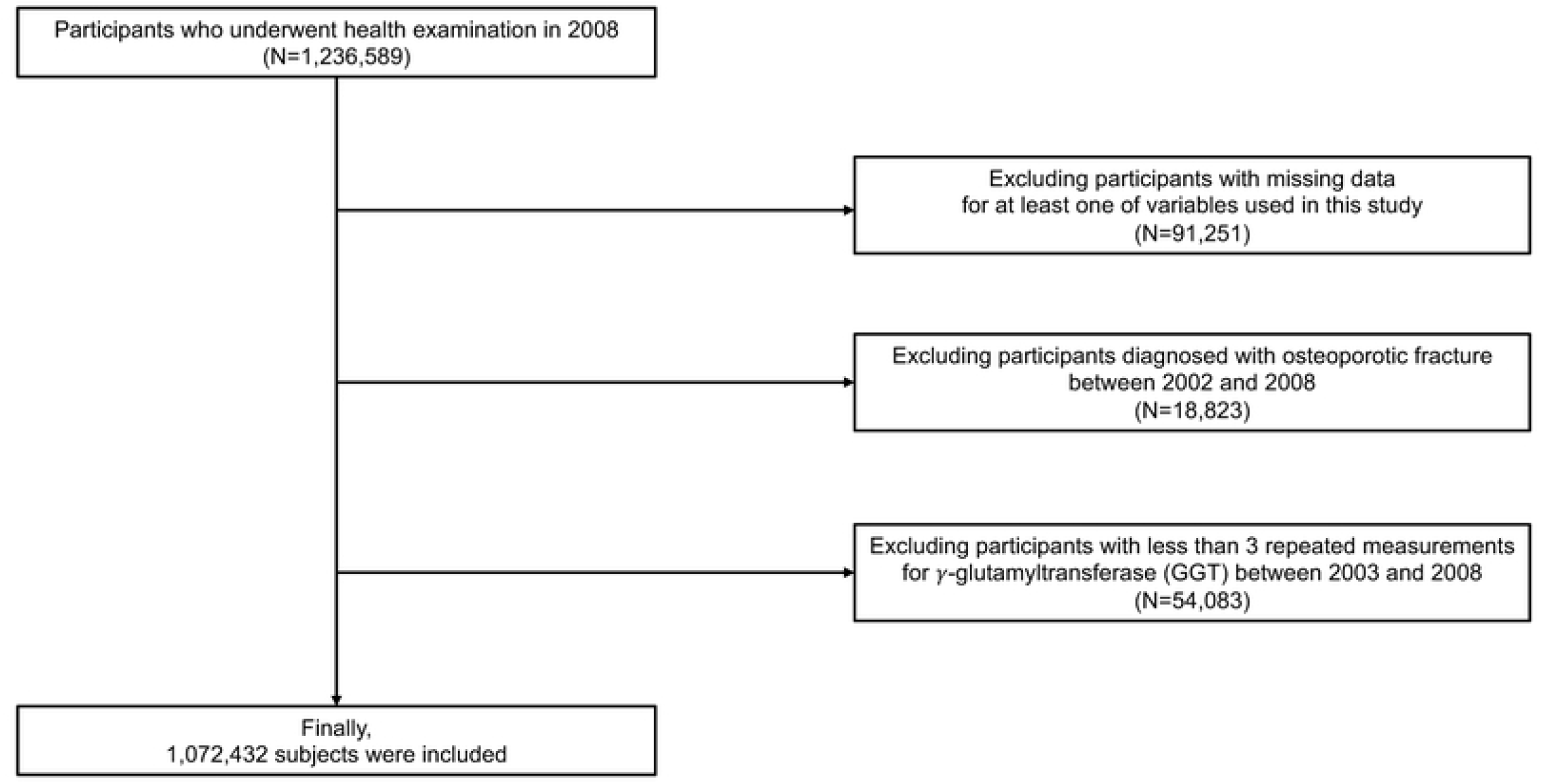
Flowchart of participant enrollment.

### Definition and variables

The index date was determined as the day of the health checkup, on which the following baseline characteristics were collected: age, sex, body mass index (BMI), and household income. Information on smoking habits (none, former, and current), alcohol consumption (frequency per week), and regular exercise (frequency per week) was obtained by questionnaires. Comorbidities were defined according to the following criteria between January 2002 and the index date. Hypertension was defined as satisfying one of the following criteria: 1) at least one claim of associated diagnostic codes (International Classification of Diseases, Tenth Revision (ICD)-10 I10–15) with prescription of an antihypertensive agent, 2) two or more claims of diagnostic codes ICD-10 E11–14, 3) systolic/diastolic blood pressure ≥140/90 mmHg, or 4) self-reported hypertension in the questionnaire. Diabetes mellitus was defined as satisfying one of the following criteria: 1) at least one claim of related diagnostic codes (ICD-10 E11–14) with prescription of an antidiabetic agent, 2) two or more claims of diagnostic codes ICD-10 E11–14, 3) fasting serum glucose level ≥ 7.0 mmol/L, or 4) self-reported diabetes mellitus in the questionnaire. Dyslipidemia was defined as satisfying one of the following criteria: 1) at least one claim of diagnostic codes (ICD-10 E78) with prescription of a dyslipidemia-related agent, 2) two or more claims of diagnostic codes (ICD-10 E78), and 3) total cholesterol ≥ 240 mg/dL. Stroke was defined as two or more claims of diagnostic code (ICD-10 I60–64). Atrial fibrillation was defined as two or more claims of diagnostic code (ICD-10 I48). Renal disease was defined as two or more claims of diagnostic codes (ICD 10 N17-19, I12-13, E082, E102, E112, E132) or estimated glomerular filtration rate less than 60 mL/min/1.73m^2^. Cancer was defined as at least one admission or at least three outpatient claims of diagnostic code (ICD-10 C00–97) with a specific registration code of ‘V027’ or ‘V193–4.’(14)

### Definition of GGT variability

GGT variability was defined as intraindividual change in GGT value in data of health checkups conducted in the 6 years before the index year (2009). Blood tests after at least 8 hours of fasting were performed in laboratories certified by the government, and the results were registered in the national database. Variability indexes were as follows: (1) coefficient of variation (CV), (2) standard deviation (SD), and (3) variability independent of the mean (VIM). The VIM was calculated as 100 x SD/Mean^beta^, where beta is the regression coefficient, on the basis of the natural logarithm of the SD over the natural logarithm of the mean.(15)

### Study outcomes

The primary outcome was occurrence of osteoporotic fractures, defined as two or more claims of one of the following ICD-10 codes 1) vertebral fractures (S22.0, S22.1, S32.0, S32.7, T08, M48.4, M48.5, M49.5), 2) hip fractures (S72.0, S72.1), distal radius fractures (S52.5, S52.6), or humerus fractures (S42.2, S42.3) based on the osteoporotic fracture fact sheet from the Korean Society of Bone and Mineral Research.(8) The follow-up period was calculated as the time between the index date and the time of osteoporotic fractures, the death of the subject, or December 2020, whichever occurred first.

### Statistical analysis

Categorical variables were presented as numbers (percentages) and continuous variables as means ± SDs. For comparisons among groups, categorical variables were analyzed using the chi-square test and continuous variables using the analysis of variance test. Restricted cubic splines were assessed to confirm the possibility of a non-linear association between GGT variability and osteoporotic fracture risk, and all GGT variability showed a positive linear association.(16) Kaplan-Meier survival curves were used to investigate the association between quartile of GGT variability and risk of osteoporotic fracture based on the log-rank test. To estimate the incidence of osteoporotic fractures, the total number of events was divided by the sum of person-years. To determine the risk of fracture according to quartile of GGT variability, Cox proportional hazard regression analysis was used, and hazard ratio (HR) with 95% confidence interval (CI) were presented. The multivariable regression model was performed with adjustment for age, sex, BMI, household income, alcohol consumption, smoking status, regular physical activity, and comorbidities (hypertension, diabetes mellitus, dyslipidemia, stroke, atrial fibrillation, cancer, and renal disease). The assumption of the proportionality of hazards was tested using Shoenfeld’s residuals. No departure from the proportional hazards assumption was detected. Subgroup analysis was performed with each kind of osteoporotic fracture (vertebral, hip, distal radius, and humerus fractures). For sensitivity analysis, (1) the mean GGT level was further adjusted in multivariable analysis, and (2) participants with osteoporotic fractures within 1 year from the index date were excluded to minimize the possibility of reverse causality. Cox regression analyses for interactions between osteoporotic fractures and the subgroups depending on covariates were performed among the quartiles of GGT variability based on the CV. Statistical Analysis System software (SAS version 9.2, SAS Institute, Cary, NC) was used for statistical analyses, and a *P*-value < 0.05 was considered to be statistically significant.

## Results

During the median of 12.3 years (interquartile range 12.1 – 12.6), osteoporotic fractures occurred in 49,677 (4.6%) patients. Regarding the sites of fracture, 24,190 participants had vertebral fractures, 3,843 had hip fractures, 21,191 had distal radius fractures, and 3,910 had humerus fractures. GGT level was measured a total of 5,647,306 times in this study (3 times in111,538 patients, 4 times in 131,344, 5 times in 189,984, and 6 times in 639,566). Table 1 demonstrates the results of comparative analysis of the groups according to quartile of GGT variability based on CV. Participants with higher quartile of GGT variability were more likely to be men and older and had higher frequencies of hypertension, diabetes mellitus, dyslipidemia, stroke, atrial fibrillation, renal disease, and cancer.

**Table 1.** Baseline characteristics of subjects according to gamma-glutamyl transferase variability.

| Variable | Total | Q1 | Q2 | Q3 | Q4 | P-value |
| --- | --- | --- | --- | --- | --- | --- |
| Number of participants (%) | 1072432 | 268078 (25.0) | 268138 (25.0) | 268108 (25.0) | 268108 (25.0) |  |
| Age, years | 43.71±10.05 | 43.72±10.17 | 43.53±9.85 | 43.49±9.87 | 44.13±10.29 | <.001 |
| Sex |  |  |  |  |  | <.001 |
| Male | 828272 (77.2) | 196733 (73.4) | 206802 (77.1) | 211617 (78.9) | 213120 (79.5) |  |
| Female | 244160 (22.8) | 71345 (26.6) | 61336 (22.9) | 56491 (21.1) | 54988 (20.5) |  |
| Body mass index (kg/m<sup>2</sup>) | 23.77±3.02 | 23.27±2.97 | 23.67±2.99 | 23.97±3.00 | 24.16±3.02 | <.001 |
| Household income |  |  |  |  |  | <.001 |
| Q1, lowest | 155494 (14.5) | 39482 (14.7) | 36473 (13.6) | 36904 (13.8) | 42635 (15.9) |  |
| Q2 | 334651 (31.2) | 83523 (31.2) | 81188 (30.3) | 82429 (30.7) | 87511 (32.6) |  |
| Q3 | 392146 (36.6) | 96297 (35.9) | 100120 (37.3) | 100084 (37.3) | 95645 (35.7) |  |
| Q4, highest | 190141 (17.7) | 48776 (18.2) | 50357 (18.8) | 48691 (18.2) | 42317 (15.8) |  |
| Smoking status |  |  |  |  |  | <.001 |
| Never | 549519 (51.2) | 149146 (55.6) | 138848 (51.8) | 132412 (49.4) | 129113 (48.2) |  |
| Former | 159648 (14.9) | 37303 (13.9) | 40208 (15.0) | 40961 (15.3) | 41176 (15.4) |  |
| Current | 363265 (33.9) | 81629 (30.5) | 89082 (33.2) | 94735 (35.3) | 97819 (36.5) |  |
| Alcohol consumption (days/week) |  |  |  |  |  | <.001 |
| None | 667082 (62.2) | 183677 (68.5) | 171397 (63.9) | 160841 (60.0) | 151167 (56.4) |  |
| 1-4 | 386503 (36.0) | 81411 (30.4) | 93046 (34.7) | 102534 (38.2) | 109512 (40.9) |  |
| ≥ 5 | 18847 (1.8) | 2990 (1.1) | 3695 (1.4) | 4733 (1.8) | 7429 (2.8) |  |
| Regular physical activity (days/week) |  |  |  |  |  | <.001 |
| None | 461467 (43.0) | 118316 (44.1) | 114913 (42.9) | 113712 (42.4) | 114526 (42.7) |  |
| 1-4 | 543876 (50.7) | 133784 (49.9) | 136839 (51.0) | 137684 (51.4) | 135569 (50.6) |  |
| ≥ 5 | 67089 (6.3) | 15978 (6.0) | 16386 (6.1) | 16712 (6.2) | 18013 (6.7) |  |
| Comorbidities |  |  |  |  |  |  |
| Hypertension | 210665 (19.6) | 43704 (16.3) | 48487 (18.1) | 53720 (20.0) | 64754 (24.2) | <.001 |
| Diabetes mellitus | 106262 (9.9) | 20469 (7.6) | 23043 (8.6) | 26972 (10.1) | 35778 (13.3) | <.001 |
| Dyslipidemia | 207890 (19.4) | 41836 (15.6) | 47363 (17.7) | 53532 (20.0) | 65159 (24.3) | <.001 |
| Stroke | 10458 (1.0) | 2102 (0.8) | 2212 (0.8) | 2514 (0.9) | 3630 (1.4) | <.001 |
| Atrial fibrillation | 4202 (0.4) | 822 (0.3) | 860 (0.3) | 951 (0.4) | 1569 (0.6) | <.001 |
| Renal disease | 12679 (1.2) | 2459 (0.9) | 2726 (1.0) | 3167 (1.2) | 4327 (1.6) | <.001 |
| Cancer | 23611 (2.2) | 5027 (1.9) | 5129 (1.9) | 5516 (2.1) | 7939 (3.0) | <.001 |
| Aspartate aminotransferase (U/L) | 25.14±12.41 | 22.94±9.08 | 24.02±10.24 | 25.41±15.77 | 29.82±32.65 | <.0001 |
| Alanine aminotransferase (U/L) | 26.12±17.42 | 22.35±13.79 | 24.83±15.85 | 27.60±19.91 | 34.10±41.54 | <.0001 |
| Mean gamma-glutamyl transferase (U/L) | 37.97±37.94 | 27.10±23.29 | 32.23±28.82 | 38.54±35.08 | 54.00±52.40 | <.001 |
| Gamma-glutamyl transferase variability |  |  |  |  |  |  |
| CV (%) | 26.85±16.15 | 11.85±3.20 | 19.83±2.04 | 27.83±2.77 | 47.91±17.38 | <.001 |
| SD | 12.00±20.61 | 3.26±3.09 | 6.42±5.88 | 10.79±10.07 | 27.53±34.69 | <.001 |
| VIM (%) | 10.51±5.86 | 4.93±1.39 | 8.08±1.14 | 11.06±1.61 | 17.97±6.20 | <.001 |

P-value by Chi-square test. Data are expressed as the mean ± SD, or n (%).
Q, quartile; CV, coefficient of variation; SD, standard deviation; VIM, variability independent of the mean.

Kaplan-Meier survival curves of freedom from osteoporotic fracture are shown in Fig 2 according to GGT variability based on CV. The risk for incident osteoporotic fracture was higher in higher quartiles of GGT variability (*P* < 0.001). In multivariable analysis, quartile of GGT variability based on CV positively correlated with the occurrence of osteoporotic fracture (adjusted HR [highest quartile compared with the lowest quartile] 1.15, 95% CI 1.12-1.18, *P* < 0.001). This trend was consistent even when GGT variability was applied to SD (adjusted HR [highest quartile compared with the lowest quartile] 1.22, 95% CI 1.19-1.25, *P* < 0.001) and VIM (adjusted HR [highest quartile compared with the lowest quartile] 1.12, 95% CI 1.09-1.15, *P* < 0.001), and the results were consistent after adjustment of mean GGT level (Table 2, Supplementary Table). When divided into deciles of GGT variability for any parameter, the risk of fracture was significantly higher in the highest 4 deciles compared to the lowest decile, and a significant trend was confirmed with an increase in the risk of fracture as decile increased (Supplementary Table 2). In addition, the association between GGT variability and osteoporotic fracture was consistently observed in landmark analysis using a start time of 1 year after the index date (Supplementary Table 3).

**Table 2.**
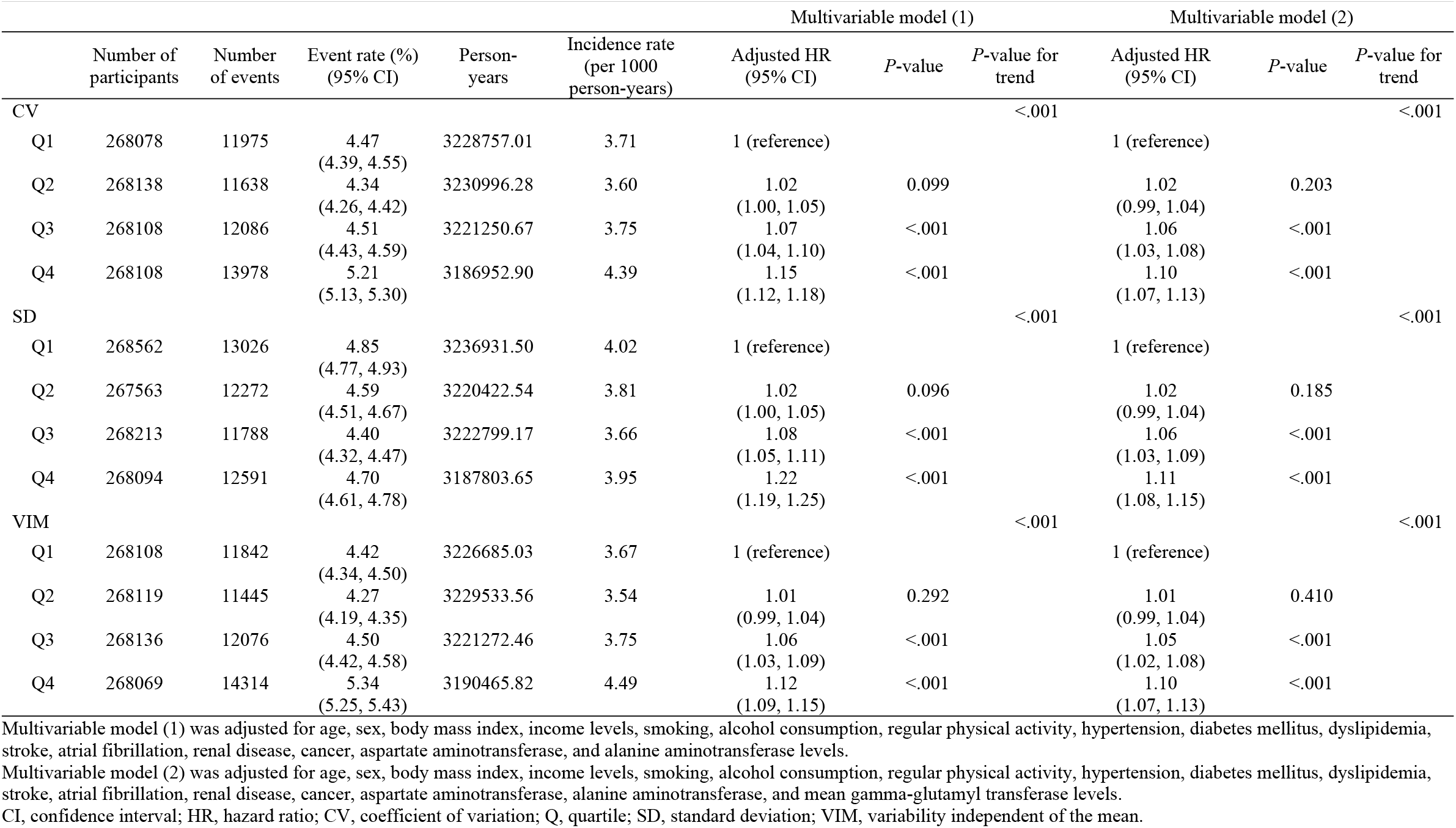
The risk for the occurrence of osteoporotic fractures according to quartiles of gamma-glutamyl transferase variability.

**Fig 2.**
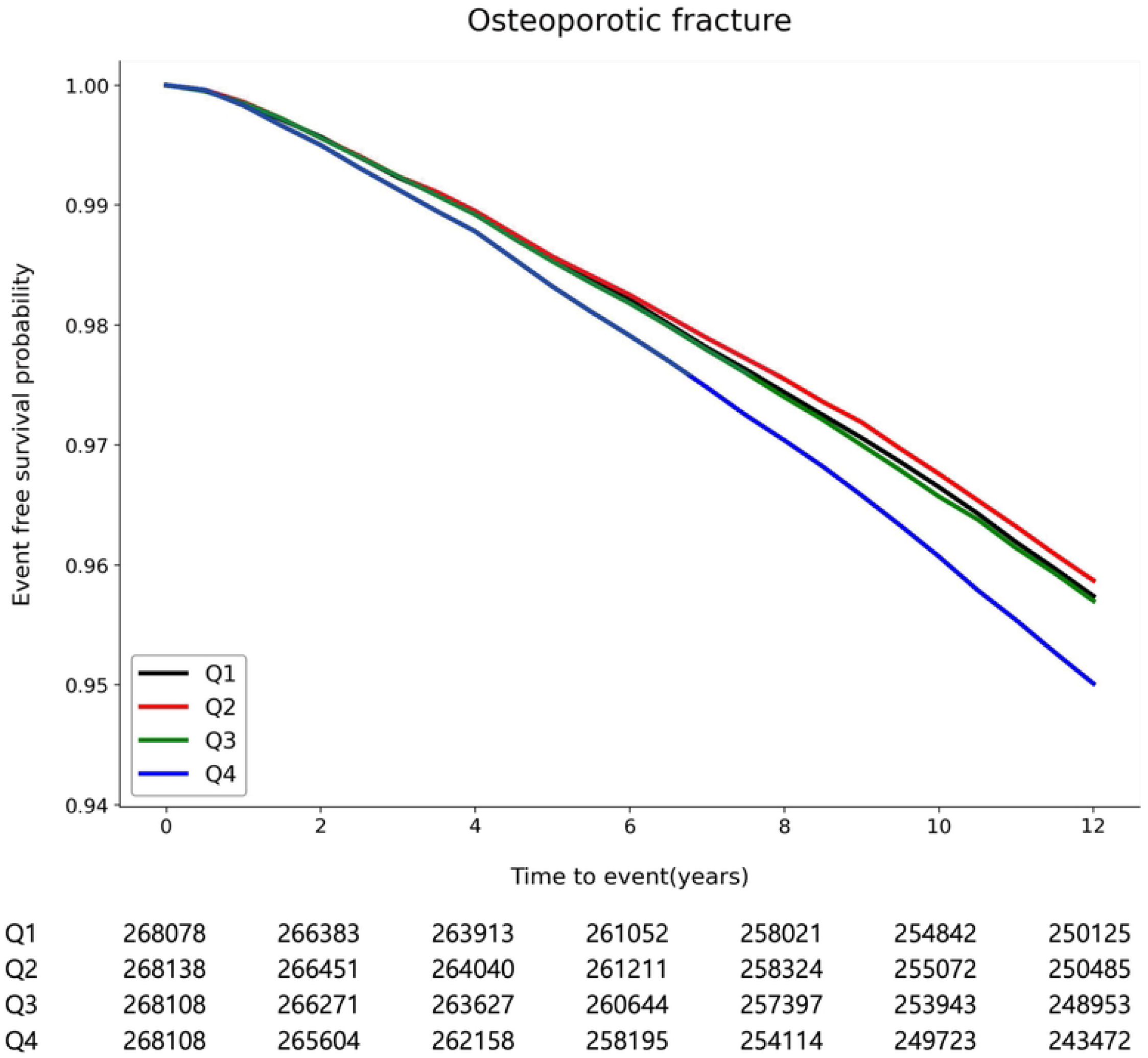
Kaplan-Meier survival curves for incident osteoporotic fracture according to gamma-glutamyl transferase variability based on the coefficient of variation. Q, quartile.

In subgroup analyses testing *P*-value for interaction, significant interaction effects regarding the presence of osteoporotic fractures were observed for sex and alcohol consumption, with a prominent association in women compared to men (adjusted HR 1.19, 95% CI 1.15-1.23 vs. adjusted HR 1.08, 95% CI 1.04-1.12, *P* = 0.035) and individuals with alcohol use ≥5 or 1-4 days/week compared to those with <1 day/week (adjusted HR 1.28, 95% CI 1.09-1.51 vs. adjusted HR 1.22, 95% CI 1.16-1.28 vs. adjusted HR 1.11, 95% CI 1.08-1.15, *P* = 0.027) (Supplementary Table 4).

In further analyses by type of osteoporotic fracture, the third and highest quartiles of GGT variability were significantly associated with risk of vertebral, hip, distal radius, and humerus fracture compared to the lowest quartiles of GGT variability regardless of variability parameter except between GGT variability based on VIM and risk of distal radius fracture (Supplementary Tables 5-8, Supplementary Fig 1).

## Discussion

The key findings of our study are that high GGT variability is associated with an increased risk of osteoporotic fracture after adjustment of potential confounders, including age, sex, BMI, alcohol consumption, physical exercise, and comorbidities. Moreover, these associations were consistent regardless of the location of fracture (vertebral, hip, distal radius, and humerus fracture) or the variability parameter (CV, SD, and VIM). In subgroup analysis, more prominent association was observed in women than men and in heavy drinkers than non-drinkers. The effect of GGT variability may be emphasized in these subgroups as woman and heavy drinking are significant risk factors for osteoporotic fractures.

Although not a large number of studies has been conducted, results have been consistent regarding serum GGT level and risk of osteoporosis represented by bone mass. A previous single clinic-based cross-sectional study demonstrated that increased GGT level was correlated with decreased BMD level measured using dual-energy X-ray absorptiometry after adjustment of confounding variables.(17) In a population-based cross-sectional study conducted in Korea, GGT level had a more distinct negative relationship with BMD at multiple bony sites compared to other liver enzymes such as aspartate transaminase and alanine transaminase, and its effect persisted after adjustment of age-sex interactions.(18)

While serum GGT level is clinically used as a biomarker indicating hepatobiliary injury or chronic alcohol consumption, one of the major physiologic roles of GGT attached to the cell membrane is to metabolize extracellular glutathione and enable the transfer of amino acids into the cell for re-synthesis of glutathione.(19) On the other hand, increased GGT activity contributes to prooxidant activity through free radical formation, lipid peroxidation, and mutagenesis, which occurs at the outer surface of the cell membrane.(20) The prooxidant and proinflammatory activities caused by increased GGT are believed to play a role in the pathogenesis of various human diseases, and GGT has been studied for its efficacy as a predictive biomarker of atherosclerosis, heart failure, metabolic syndrome, non-alcoholic liver diseases, chronic kidney disease, cancers, and fractures.(19,21,22)

Oxidative stress, which is associated with increased GGT activity, has been extensively studied in terms of bone remodeling and suggested to contribute to pathological changes in mineralized tissue. Reactive oxygen species are involved in osteoclast formation and subsequent bone resorption in mouse marrow culture cells(23) and also in human bone marrow stromal cells via expression of receptor activator of nuclear factor-kappa B ligand (RANKL) and macrophage colony-stimulating factor.(24) Moreover, oxidative stress is suggested to inhibit the differentiation of osteoblasts by activation of extracellular signal-regulated kinase (ERK) and ERK-dependent NF-κB signaling pathways.(25) Decreased bone mineral density at various sites was confirmed to be related to increased oxidative biomarkers in humans.(26,27) The relationship between GGT and bone homeostasis is thought to be mainly mediated by osteoclastic activation. Extracellular GGT protein induces osteoclastogenesis in mouse bone marrow cells through the expression of RANKL and Toll-like receptor 4.(28,29) These findings were confirmed by observation of enhanced bone resorption and subsequent osteoporosis-like conditions in transgenic mice with overexpression of GGT.(30)

Several epidemiologic studies have also found a relationship between GGT level and fracture risk, and it seems clear and consistent that there is a positive correlation, in particular with hip fracture risk. A single clinic-based longitudinal study conducted in Korea was the first to report that high baseline serum GGT level in men over 50 years of age may be useful as a biomarker for future osteoporotic fracture risk, regardless of drinking habits.(31)

Large population-based cohort studies have also recently confirmed a significant association between GGT level and hip fracture risk.(32,33) However, there has been no previous study using GGT variability as a biomarker for risk assessment of osteoporotic fractures. The high variability of biomarkers has been attracting attention as a new factor predicting poor clinical outcomes of various diseases, and high GGT variability was identified as a predictor of increased risk of myocardial infarction, stroke, dementia, chronic kidney disease, and mortality.(34–36)

We acknowledge several limitations in this study. First, additional confounding factors affecting the development of osteoporotic fractures may not have been considered. Second, since this study was conducted with the Korean population, application to other races or ethnic groups may lead to errors. Third, it is difficult to present a clear cause of GGT variability. Fourth, our retrospective observational study design cannot suggest a causal relationship. On the other hand, there are strengths of this study as well. We used large national representative data with a long follow up to elucidate the effect of GGT variability on osteoporotic fracture occurrence. Our large-scale epidemiologic study provides new perspectives supporting the benefits of maintaining GGT level for prevention of osteoporotic fractures.

## Conclusion

This nationwide population-based cohort study conducted in Korea demonstrated that an increase in GGT variability is associated with an increased risk of osteoporotic fractures, regardless of site. Maintaining stable GGT level may help reduce the risk of osteoporotic fractures.

## Data Availability

The data used in this study are available in the National Health Insurance Service-National Health Screening Cohort (NHIS-HEALS) database, but restrictions apply to public availability of these data used under license for the current study. Requests for access to the NHIS data can be made through the National Health Insurance Sharing Service homepage [http://nhiss.nhis.or.kr/bd/ab/bdaba021eng.do]. For access to the database, a completed application form, research proposal, and approval from the institutional review board should be submitted to the inquiry committee of research support in the NHIS for review.

## Authors’ contributions

DK and JHK contributed to data interpretation and drafted the manuscript. HL, IH, and YC contributed to data analysis, and visualization. TJS contributed to conception, design, data acquisition, interpretation, and critical revision of the manuscript. All authors have read and agreed to the published version of the manuscript.

## Ethical approval statement

The Institutional Review Board of Ewha Womans University College of Medicine (SEUMC 2022–07–053) approved the analysis and provided a consent waiver, as the data were anonymized and freely accessible by the NHIS for study purposes.

## Supporting information captions

Supplementary Fig 1. Kaplan-Meier survival curves for incident osteoporotic fracture at various sites according to gamma-glutamyl transferase variability based on the coefficient of variation. Q, quartile.

Supplementary Table 1. Risk factors for the occurrence of osteoporotic fractures.

Supplementary Table 2. The risk for the occurrence of osteoporotic fractures according to deciles of gamma-glutamyl transferase variability.

Supplementary Table 3. The risk for the occurrence of osteoporotic fractures according to quartiles of gamma-glutamyl transferase variability, landmark analysis using a start time of 1 year after the index date.

Supplementary Table 4. The subgroup analysis regarding gamma-glutamyl transferase variability based on the coefficient of variation and osteoporotic fractures in association with demographics or comorbidities.

Supplementary Table 5. The risk for the occurrence of vertebral fractures according to quartiles of gamma-glutamyl transferase variability.

Supplementary Table 6. The risk for the occurrence of hip fractures according to quartiles of gamma-glutamyl transferase variability.

Supplementary Table 7. The risk for the occurrence of distal radius fractures according to quartiles of gamma-glutamyl transferase variability.

Supplementary Table 8. The risk for the occurrence of humerus fractures according to quartiles of gamma-glutamyl transferase variability.

